# Tocilizumab and sarilumab alone or in combination with corticosteroids for COVID-19: A systematic review and network meta-analysis

**DOI:** 10.1101/2021.07.05.21259867

**Authors:** Dena Zeraatkar, Ellen Cusano, Juan Pablo Díaz Martinez, Anila Qasim, Sophia O. Mangala, Elena Kum, Jessica J. Bartoszko, Tahira Devji, Thomas Agoritsas, Francois Lamontagne, Bram Rochwerg, Per O Vandvik, Romina Brignardello-Petersen, Reed Siemieniuk

## Abstract

**Objective:** To compare the effects of interleukin-6 (IL-6) receptor blockers, with or without corticosteroids, on mortality in patients with COVID-19.

**Design:** Systematic review and network meta-analysis

**Data sources:** WHO COVID-19 database, a comprehensive multilingual source of global covid-19 literature, and two prospective meta-analyses

**Study selection:** Trials in which people with suspected, probable, or confirmed COVID-19 were randomized to IL-6 receptor blockers (with or without corticosteroids), corticosteroids, placebo, or standard care.

**Results:** We assessed the risk of bias of included trials using a modification of the Cochrane risk of bias tool. We performed a Bayesian fixed effect network meta-analysis and assessed the certainty of evidence using the GRADE approach.

We identified 45 eligible trials (20,650 patients), 36 (19,350 patients) of which could be included in the network meta-analysis. 27 of 36 trials were rated at high risk of bias, primarily due to lack of blinding. Tocilizumab (20 more per 1000, 15 fewer to 59 more; low certainty) and sarilumab (11 more per 1000, 38 fewer to 55 more; low certainty) alone may not reduce the risk of death. Tocilizumab, in combination with corticosteroids, probably reduces the risk of death compared to corticosteroids alone (35 fewer per 1000, 52 fewer to 18 more; moderate certainty) and sarilumab, in combination with corticosteroids, may reduce the risk of death compared to corticosteroids alone (43 fewer, 73 fewer to 12 more; low certainty). Tocilizumab and sarilumab, both in combination with corticosteroids, may have similar effects (8 more per 1000, 20 fewer to 35 more; low certainty).

**Conclusion:** IL-6 receptor blockers, when added to standard care that includes corticosteroids, in patients with severe or critical COVID-19, probably reduce mortality. Tocilizumab and sarilumab may have similar effectiveness.

**Systematic review registration:** *NA*

**What is already known on this topic?:**

- IL-6 receptor blockers have immunosuppressive effects that may be important in COVID-19 patients with immune system dysfunction and inflammation
- Corticosteroids reduce the risk of death in patients with severe or critical COVID-19

**What this study adds:**

- Our systematic review and network meta-analysis provides a comprehensive review of the evidence addressing the effects of IL-6 receptor blockers, alone or in combination with corticosteroids, in COVID-19
- IL-6 receptor blockers when added to a standard care that includes corticosteroids, in patients with severe or critical COVID-19, probably reduce mortality.
- Tocilizumab and sarilumab in combination with corticosteroids may have similar effectiveness for reducing mortality.

## Background

As of June 2021, there have been more than 180 million cumulative cases of coronavirus disease 19 (COVID-19) worldwide and nearly four million deaths (1). In an attempt to improve outcomes for patients with COVID-19, several drugs have been repurposed with varying results (2). Corticosteroids are the only medication so far to have reduced mortality in patients with severe and critical disease (2). Tocilizumab, an interleukin-6 (IL-6) receptor blocker may also reduce mortality, but whether sarilumab (another IL-6 receptor blocker) reduces mortality is uncertain.

IL-6 receptor blockers have immunosuppressive effects that may be important in COVID-19 patients with immune system dysfunction and inflammation (3–5). The RECOVERY trial found that tocilizumab reduces mortality and ventilation, particularly among patients receiving corticosteroids (6) and REMAP-CAP showed tocilizumab and sarilumab to reduce mortality and improve organ-support free days (7). Results across other trials, however, have not been consistent (8–10). A prospective meta-analysis also found that tocilizumab reduces mortality (11), but whether sarilumab reduces mortality compared to no IL-6 receptor blocker, and its effect relative to tocilizumab is unclear.

Tocilizumab is an expensive drug to which patients with COVID-19 currently have access—in settings where it is available, tocilizumab is often used in only a minority of patients who might benefit from it (12). If sarilumab is a comparable alternative to tocilizumab, it would increase availability for patients with COVID-19 who would not have otherwise have access to IL-6 receptor blocker.

Here we report a systematic review and network meta-analysis addressing the effectiveness of IL-6 receptor blockers, alone or in combination with corticosteroids, for COVID-19. This review capitalizes on the methods and data of our living systematic review and network meta-analysis of drug therapies for COVID-19 (2). This evidence synthesis is part of the BMJ Rapid Recommendations project, to inform World Health Organization (WHO) Living Guidelines on drugs for treatment of COVID-19 (13).

## Methods

A protocol of our methods is contained as a supplement to our living systematic review and network meta-analysis (SRNMA) of drug therapies for COVID-19 (2).

### Search

Our study uses the search strategy of our living SRNMA that includes daily searches in the World Health Organization (WHO) COVID-19 database—a comprehensive multilingual source of global published and preprint literature on COVID-19 (https://search.bvsalud.org/global-literature-on-novel-coronavirus-2019-ncov/). Prior to its merge with the WHO COVID-19 database on 9 October 2020, we searched the US Centers for Disease Control and Prevention (CDC) COVID-19 Research Articles Downloadable Database. The database includes, but is not limited to the following bibliographic and grey literature sources: Medline (Ovid and PubMed), PubMed Central, Embase, CAB Abstracts, Global Health, PsycInfo, Cochrane Library, Scopus, Academic Search Complete, Africa Wide Information, CINAHL, ProQuest Central, SciFinder, the Virtual Health Library, LitCovid, WHO covid-19 website, CDC covid-19 website, Eurosurveillance, China CDC Weekly, Homeland Security Digital Library, ClinicalTrials.gov, bioRxiv (preprints), medRxiv (preprints), chemRxiv (preprints), and SSRN (preprints). Our search also includes six Chinese databases: Wanfang, Chinese Biomedical Literature, China National Knowledge Infrastructure, VIP, Chinese Medical Journal Net (preprints), and ChinaXiv (preprints). A validated machine learning model facilitates efficient identification of randomized trials (14). We searched WHO information sources from 1 December 2019 to 9 June 2021 and the Chinese literature from conception of the databases to 20 February 2021.

Our search is supplemented by ongoing surveillance of living evidence retrieval services, including the Living Overview of the Evidence (L-OVE) COVID-19 platform by the Epistemonikos Foundation (https://app.iloveevidence.com/loves/5e6fdb9669c00e4ac072701d) and the Systematic and Living Map on COVID-19 Evidence by the Norwegian Institute of Public Health (https://www.fhi.no/en/qk/systematic-reviews-hta/map/).

In addition, we include data directly from trialists via two WHO-sponsored prospective meta-analyses (11, 15). We report our full strategy as a supplement to our drug therapy publication (2).

### Study selection

As part of the living SRNMA, pairs of reviewers, following calibration exercises, worked independently and in duplicate to screen titles and abstracts of search records and subsequently the full texts of records determined potentially eligible at the title and abstract screening stage and to link preprint reports with their subsequent publications based on trial registration numbers, authors, and other trial characteristics. Reviewers resolved discrepancies by discussion, and when necessary, by adjudication with a third-party reviewer.

We included preprint and peer reviewed reports of trials that compared IL-6 receptor blockers with standard care, placebo, or corticosteroids or that compared corticosteroids with standard care or placebo in patients with suspected, probable, or confirmed COVID-19. There were no restrictions on severity of illness, setting, or language of publication.

### Data collection

As part of the living SRNMA, for each eligible trial, pairs of reviewers, following training and calibration exercises, independently extracted trial characteristics (trial registration, publication status, study design), patient characteristics (country, age, sex, type of care, severity of COVID-19 symptoms), and outcomes of interest (means or medians and measures of variability for continuous outcomes and the number of participants analyzed and the number of participants who experienced an event for dichotomous outcomes) using a standardized, pilot tested data extraction form. Reviewers resolved discrepancies by discussion and, when necessary, with adjudication by a third party. We updated our data when a study preprint becomes available as a peer reviewed publication. For this review, we focus on all-cause mortality closest to 90 days.

To assess risk of bias, reviewers, following training and calibration exercises, use a revision of the Cochrane tool for assessing risk of bias in randomized trials (RoB 2.0) (16). We present our modified risk of bias tool as a supplement to our drug therapy publication (2). Reviewers resolved discrepancies by discussion and, when necessary, by adjudication with a third party.

### Analysis

We performed network meta-analysis using a Bayesian framework to compare tocilizumab with and without corticosteroids, sarilumab with and without corticosteroids, corticosteroids without IL-6 receptor blockers, and standard care or placebo. We used a plausible prior for the variance parameter and a uniform prior for the effect parameter (17). We used three Markov chains with 100,000 iterations after an initial burn-in of 10,000 and a thinning of 10. We used node splitting models to assess local incoherence and to obtain indirect estimates. All network meta-analyses were performed using the gemtc package of R version 3.6.3 (RStudio, Boston, MA) and all pairwise meta-analyses using the bayesmeta package. We produced network plots using the network map command of Stata version 17.0 (StataCorp, College Station, TX) with nodes weighted by the number of studies evaluating each treatment and edges weighted by the inverse variance of the direct estimate (18).

We present fixed-effect meta-analyses as the primary analysis and random-effects meta-analyses as sensitivity analysis because we found the estimates from random-effects to have credible intervals that were implausibly wide due to the uncertainty around the heterogeneity estimate.

We summarized the effect of interventions on mortality using odds ratios and corresponding 95% credible interval.

### Certainty of evidence

To facilitate interpretation of the results, we calculated absolute effects for mortality using data from the CDC on patients who were hospitalized for COVID-19 (19, 20). For duration of ventilation, we used baseline risks from the International Severe Acute Respiratory and Emerging Infection COVID-19 database (21).

We assessed the certainty of evidence using GRADE approach for network meta-analysis, using a minimally contextualized approach, with a null effect as the threshold of importance (22–25). The minimally contextualized approach considers only whether credible intervals include the null effect and thus does not consider whether plausible effects, captured by credible intervals, include both important and trivial effects. To evaluate certainty of no benefit (or no effect), we used a 1% risk difference threshold of the 95% credible interval for mortality. We decided on this preliminary threshold based on a survey of the authors.

Two reviewers with experience in applying the GRADE approach rated each domain for each comparison separately and resolved discrepancies by consensus. We rated the certainty for each comparison and outcome as high, moderate, low, or very low, based on considerations of risk of bias, inconsistency, indirectness, publication bias, intransitivity, incoherence (difference between direct and indirect effects), and imprecision. We rated down for risk of bias if the interpretation of the effect would change if only studies at low risk of bias would have been considered. For example, if the credible interval of the pooled effect from studies at low risk of bias would have crossed the threshold for imprecision, we rated down for risk of bias.

### Patient and public involvement

Patients were involved in outcome selection, interpretation of results, and the generation of parallel recommendations, as part of the WHO Rapid Recommendations initiative, in partnership with The BMJ and MAGIC Evidence Ecosystem Foundation.

## Results

### Study characteristics

We screened 45,854 titles and abstracts and 884 full-texts and identified 45 eligible trials, including 20,650 patients (6, 26–48). Figure 1 presents details about study selection. Twenty-one of these trials were published, four were available as preprints, and 20 were unpublished and were retrieved from two prospective meta-analyses (11, 49).

**Figure 1:**
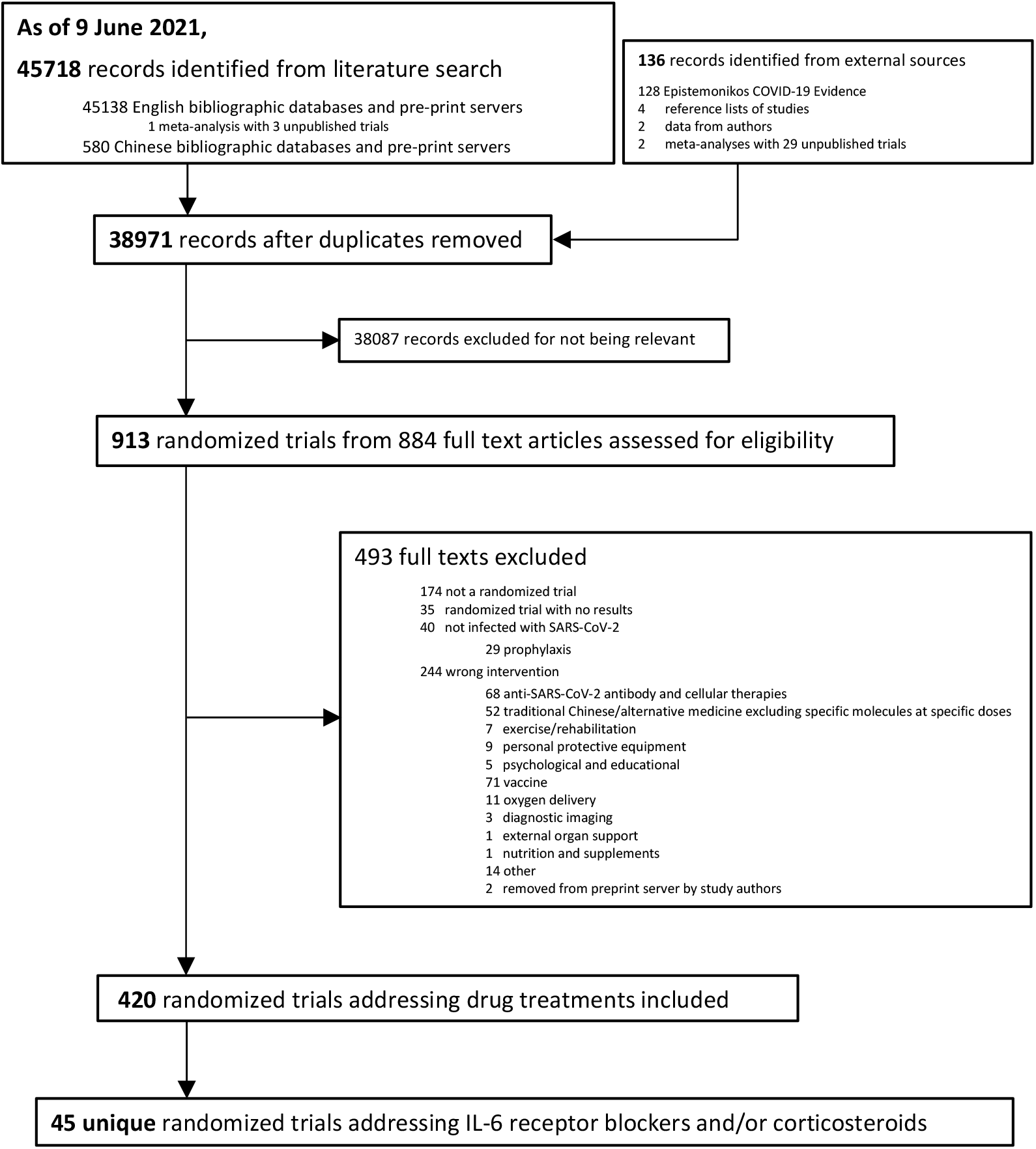
PRISMA diagram.

Table 1 presents trial characteristics. Twenty trials (7,608 patients) compared tocilizumab with standard care or placebo, (6, 26–34) seven trials (2,756 patients) sarilumab with standard care or placebo with or without corticosteroids,(35, 36) three trials (366 patients) compared IL-6 receptor blockers with corticosteroids (48), and 14 trials (8,102 patients) corticosteroids with standard care or placebo (37–47).

**Table 1:**
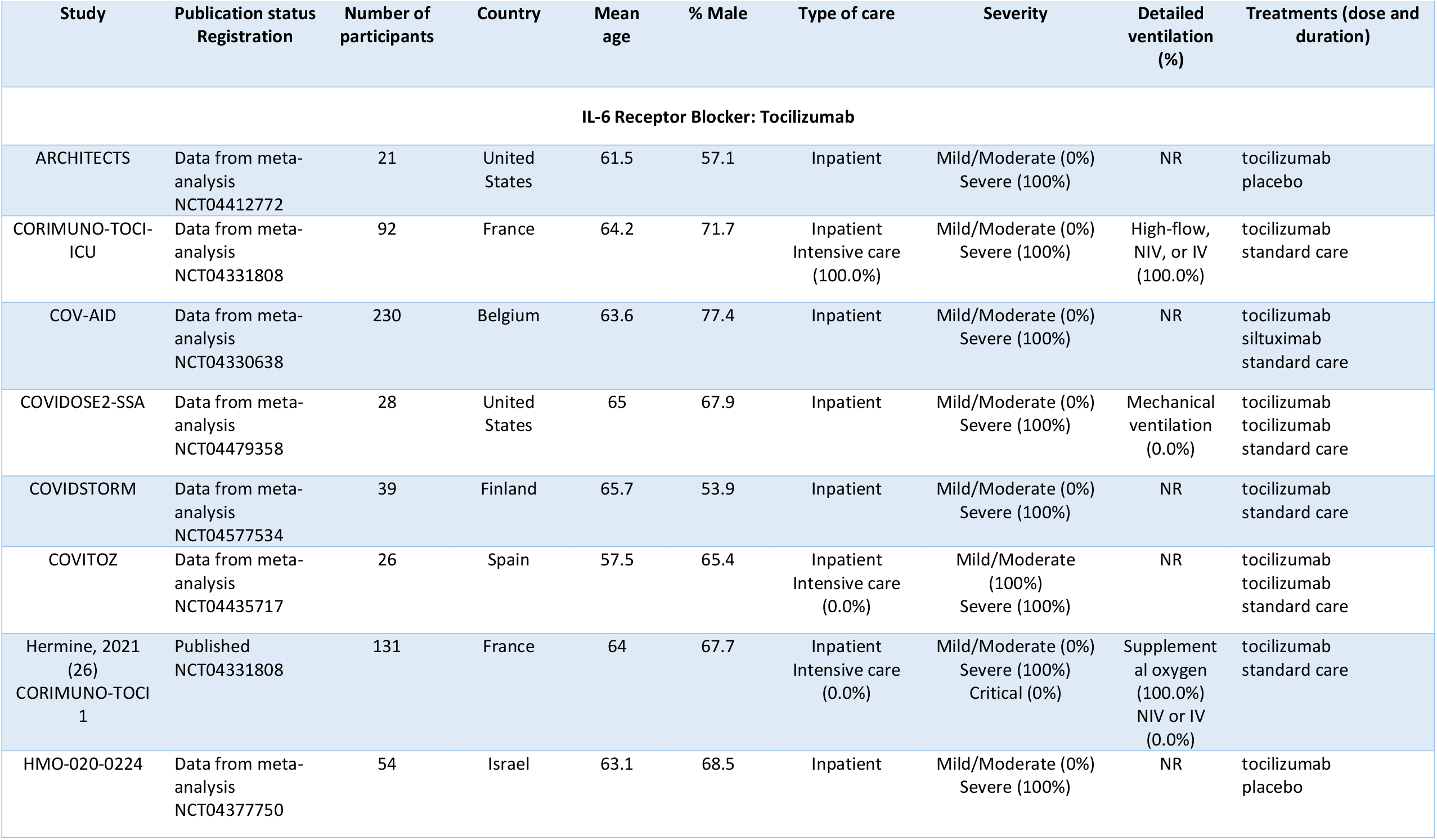
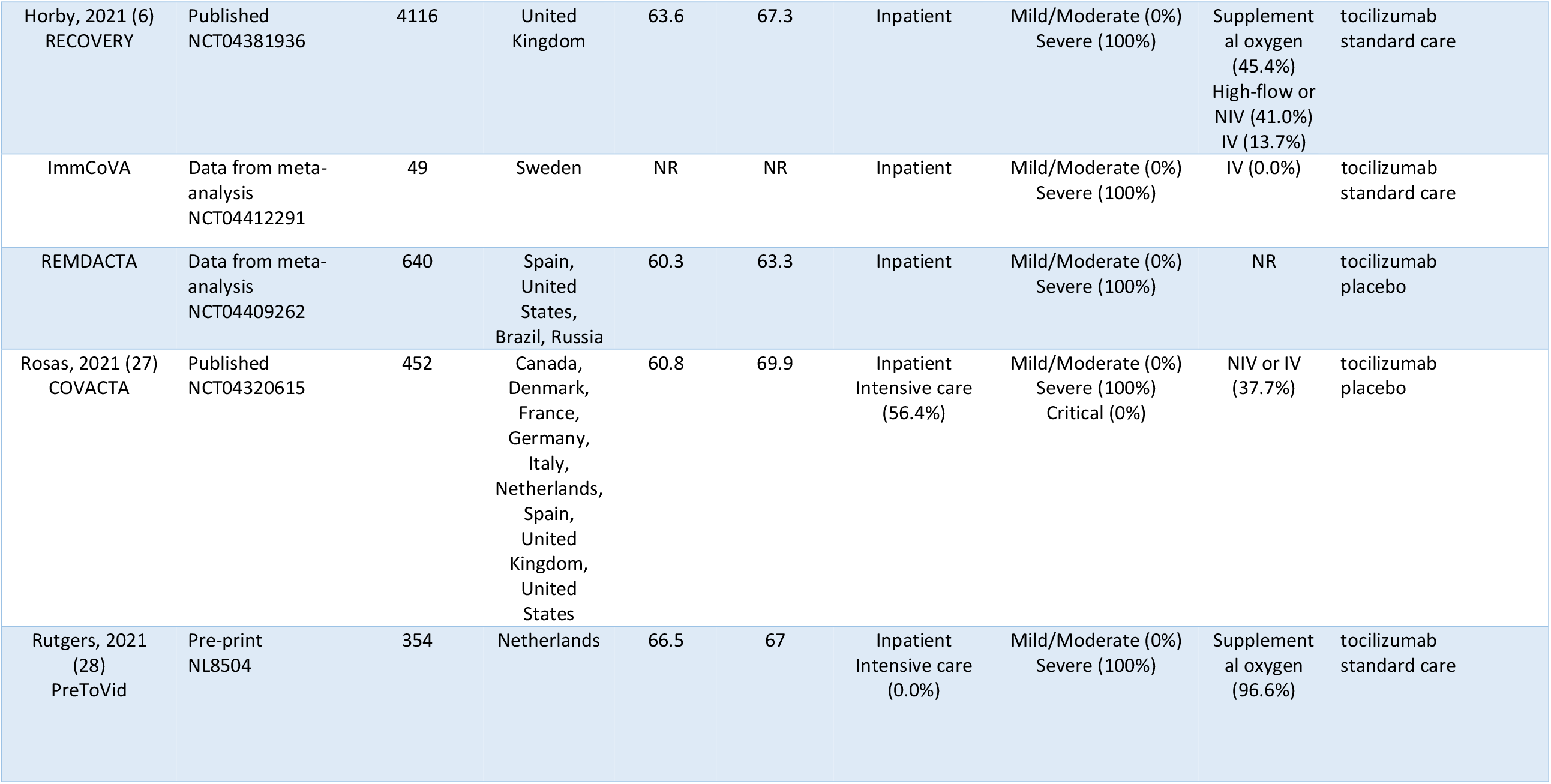
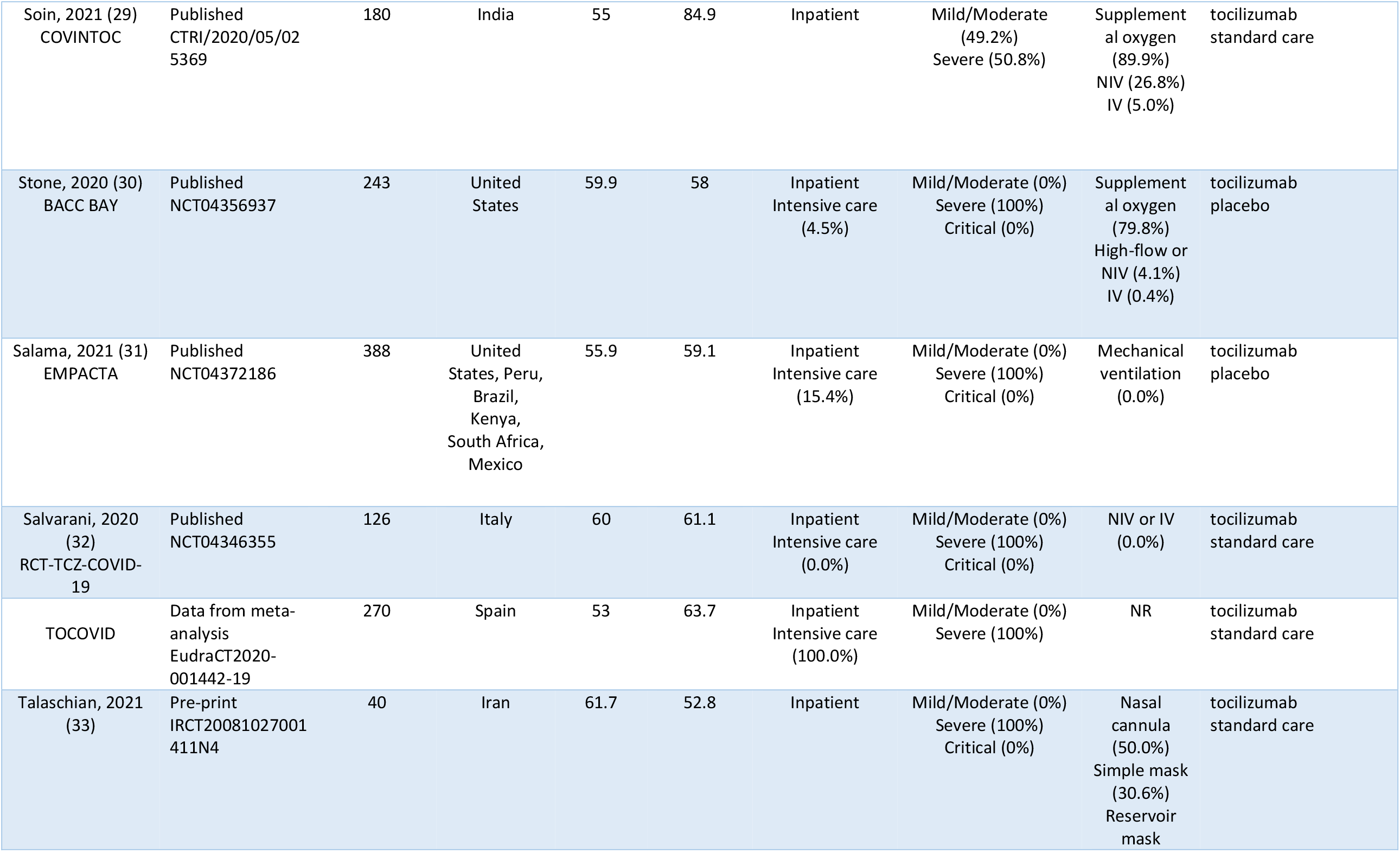
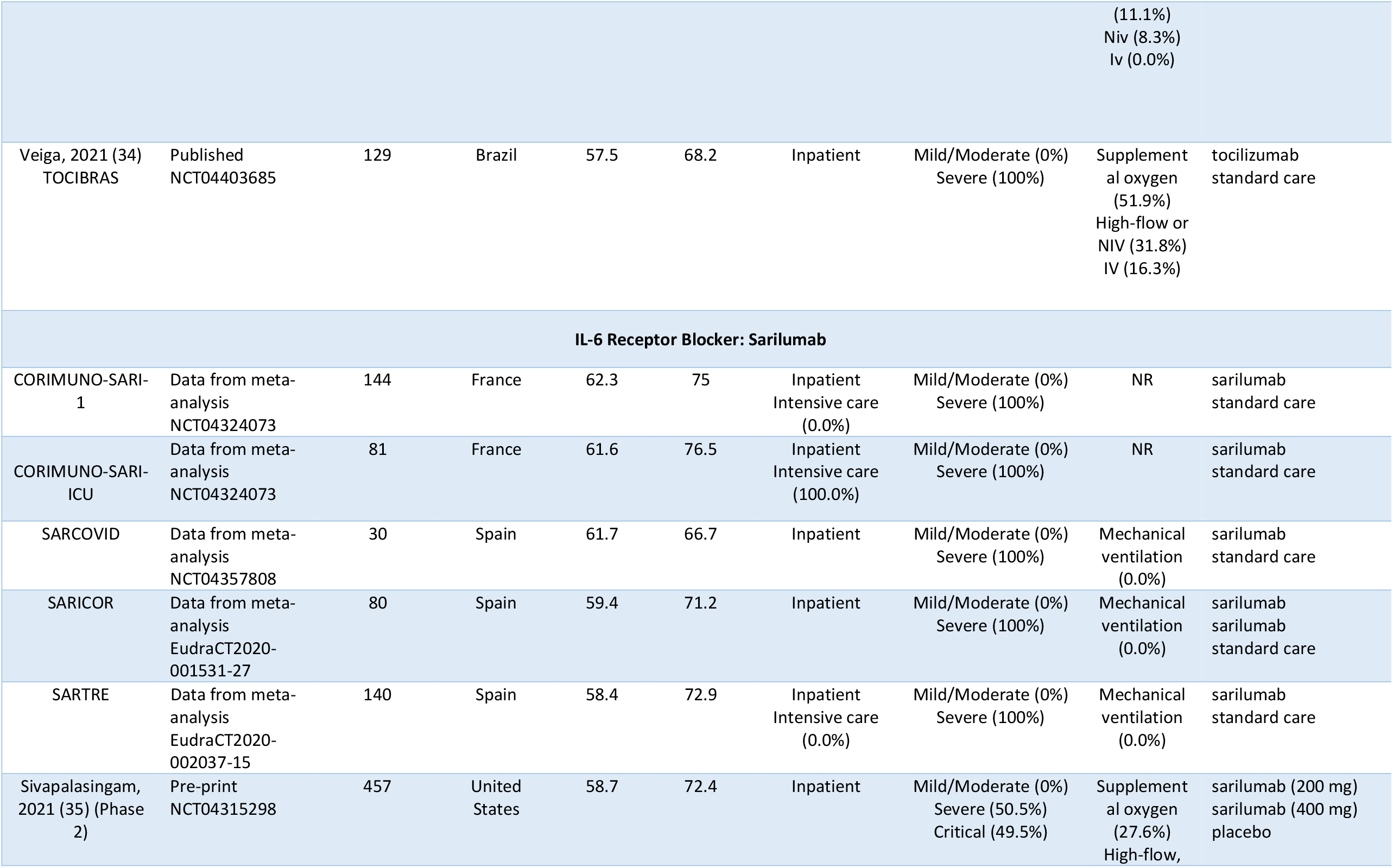
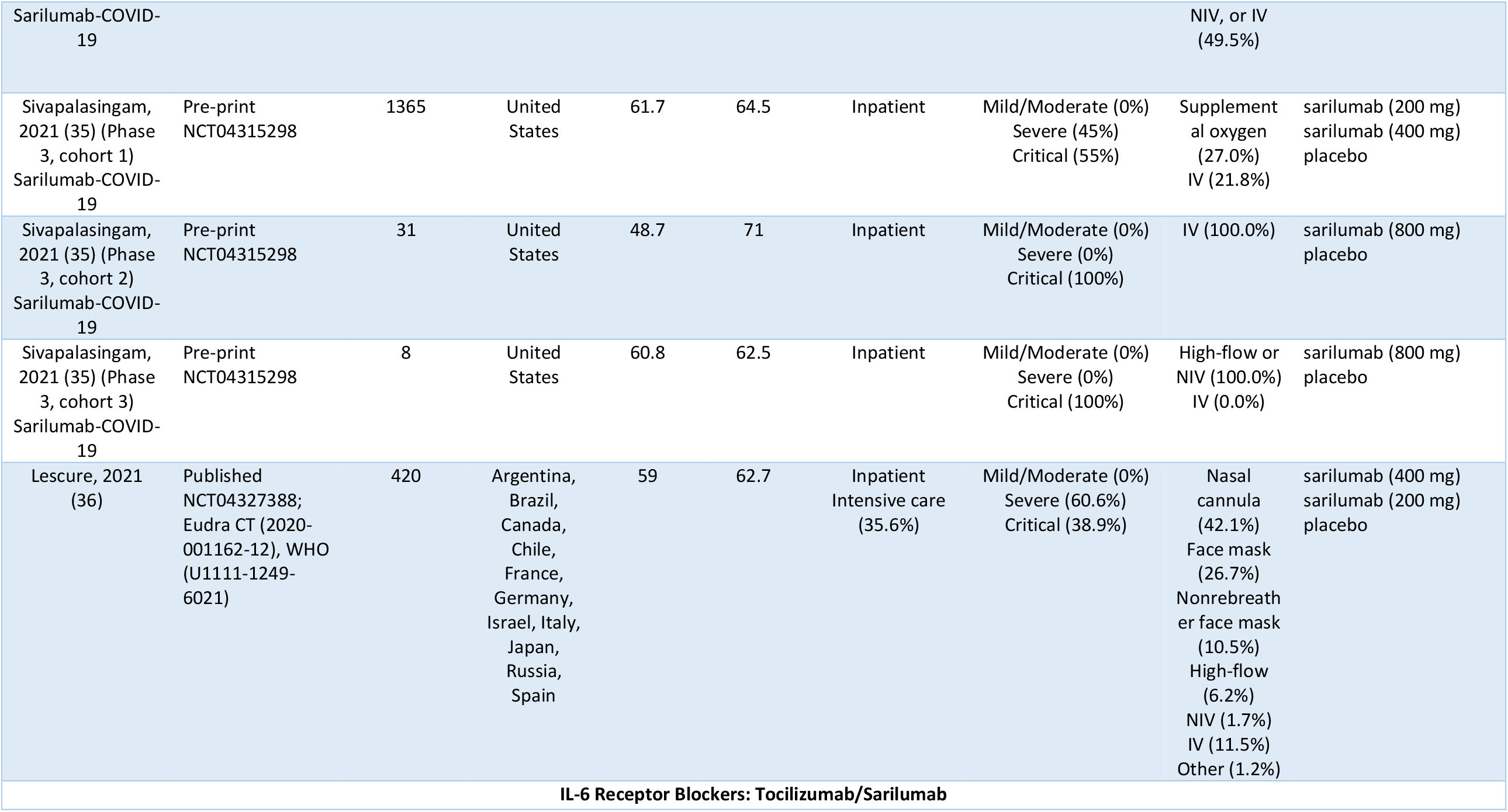
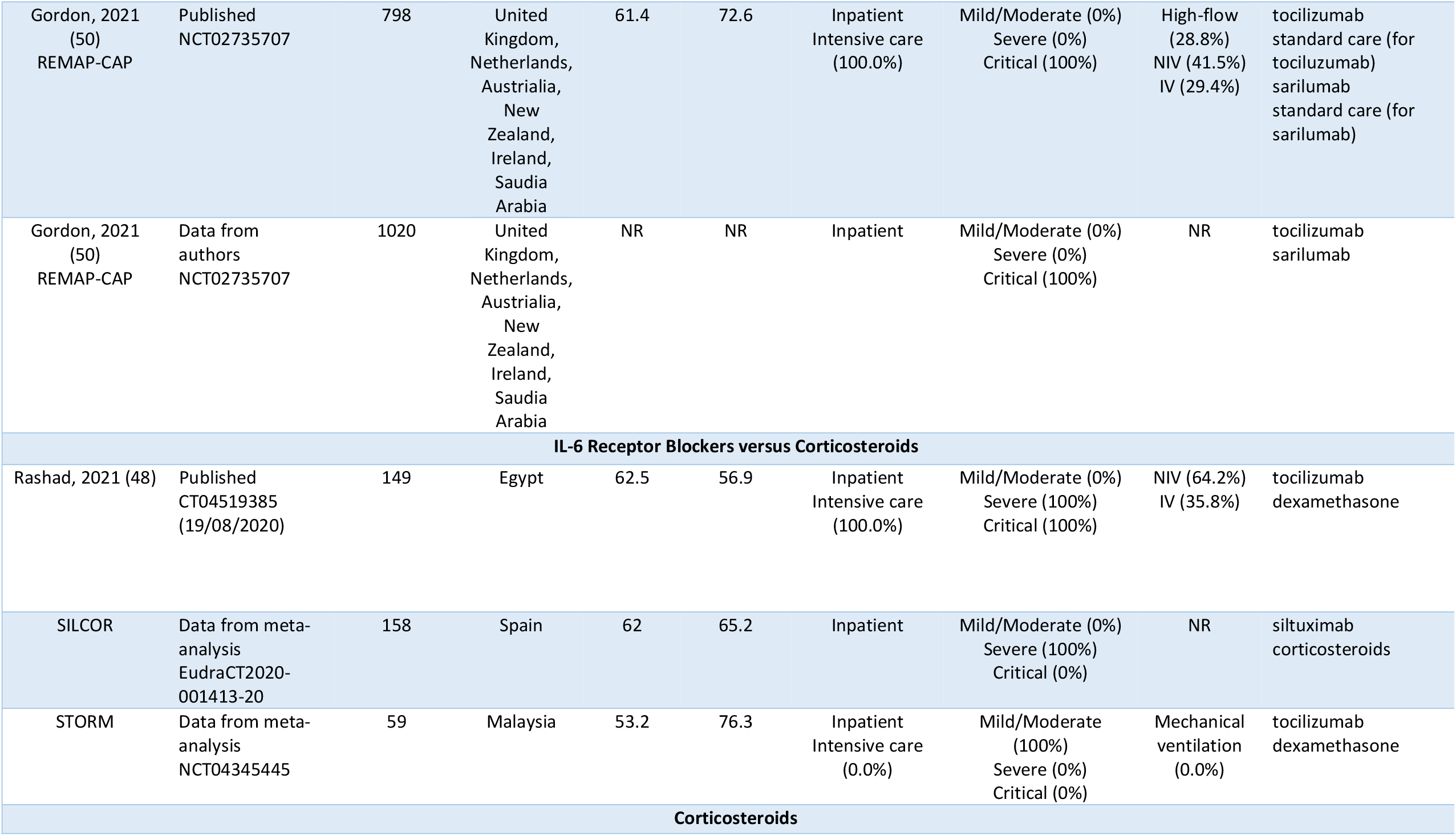
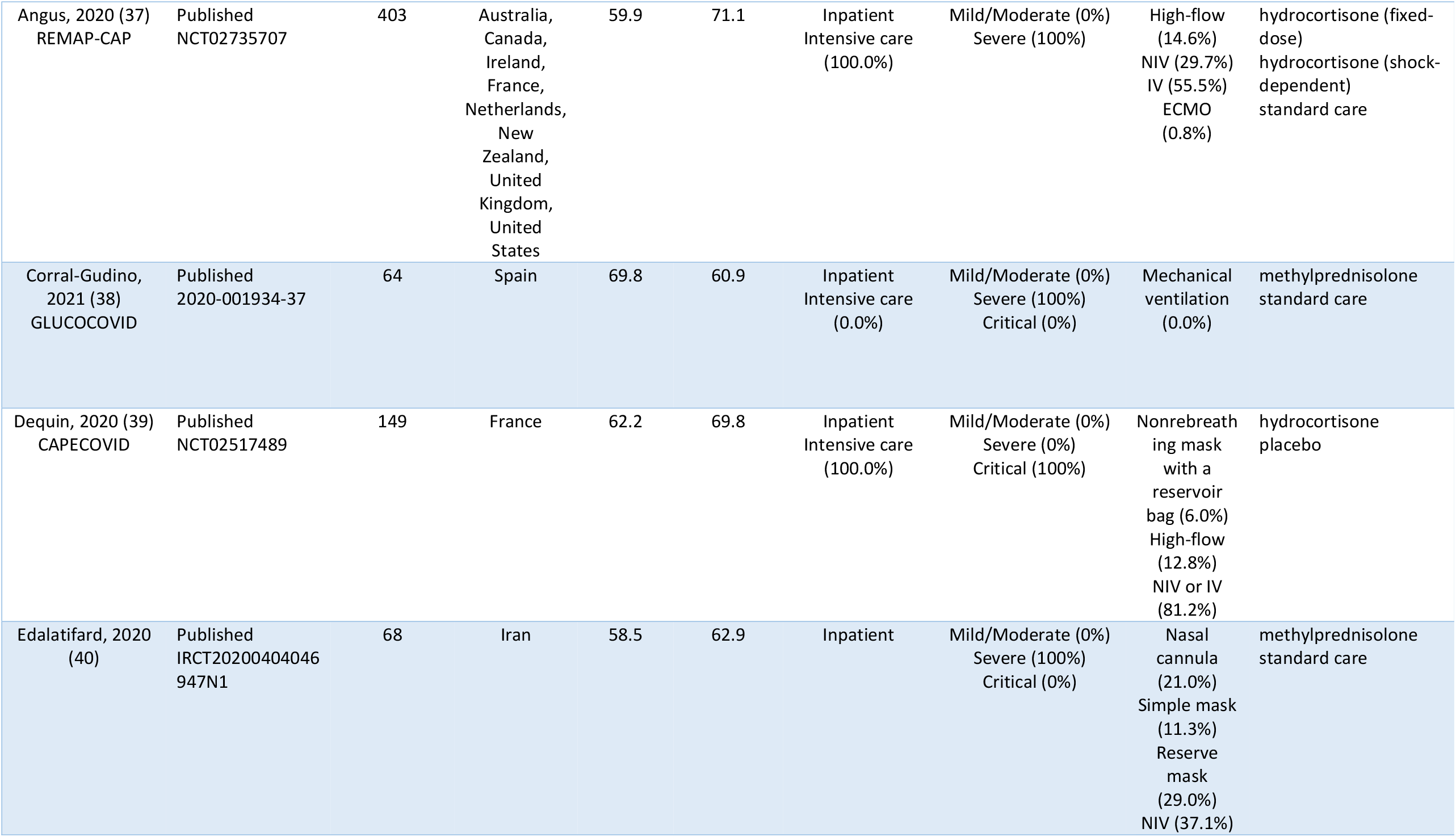
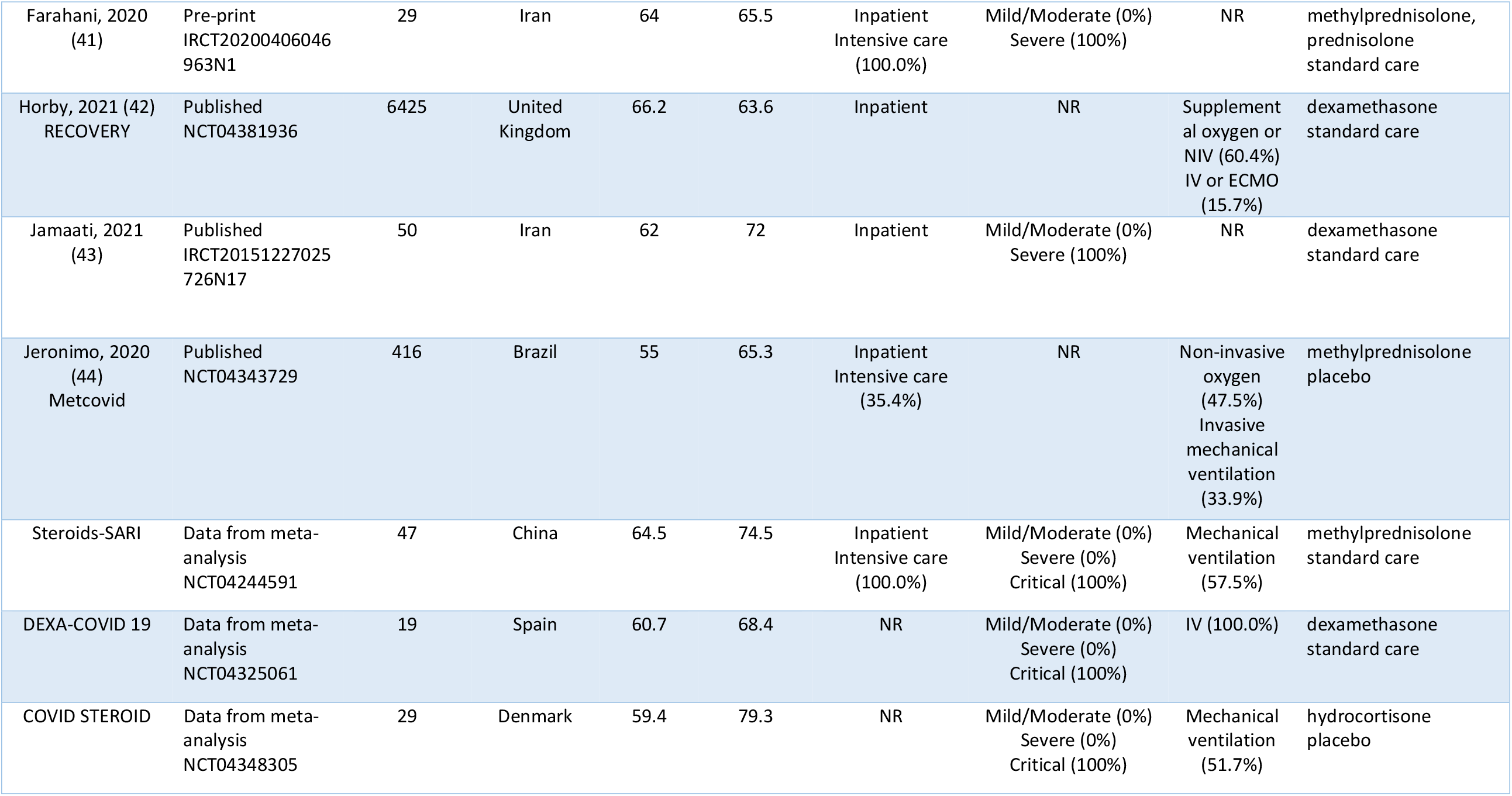
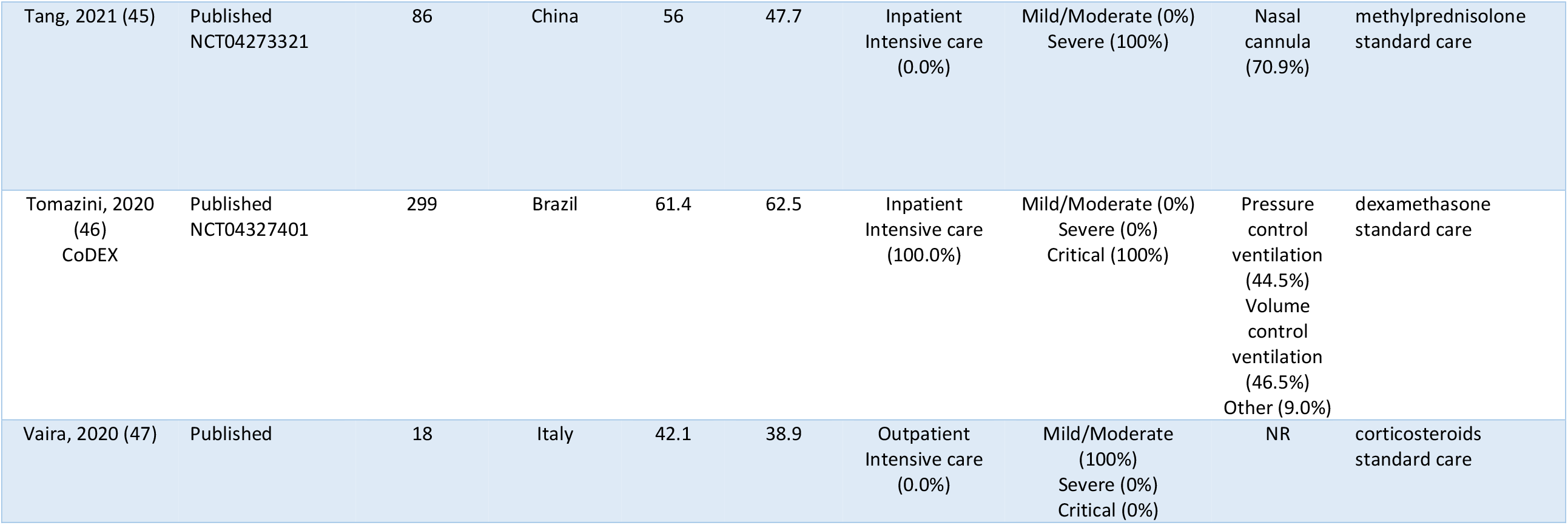
Trial Characteristics.

One trial, REMAP-CAP (7), randomized patients to tocilizumab or standard care (among centres with access to tocilizumab) or to sarilumab or standard care (among centres with access to sarilumab). Randomization to standard care was halted midway through the trial when an interim analysis showed efficacy of tocilizumab and sarilumab after which patients were randomized to either tocilizumab or sarilumab, with both groups receiving corticosteroids. We treated REMAP-CAP as three separate trials in our analyses (i.e., tocilizumab versus standard care; sarilumab versus standard care; tocilizumab versus sarilumab). We used 90-day mortality for the comparisons of tocilizumab and sarilumab with standard care and obtained data on in-hospital mortality from the investigators of the trial on the comparison of tocilizumab and sarilumab.

Another trial, Sarilumab-COVID-19, was conducted in two phases (35). Phase three underwent two protocol amendments involving patient eligibility and dose. The trial was thus treated as four separate trials.

### Patient characteristics

Table 1 presents characteristics of patients included in the trial. Trials included a median of 129 [IQR: 47 to 354] participants. The mean age of patients in trials ranged between 42.1 to 69.8. About half of all patients were recruited from the United Kingdom. All but one trial reported on inpatients. Almost all trials reported on patients with severe to critical disease and most patients were receiving some form of supplementary oxygen.

### Risk of bias

Figure 3 presents risk of bias assessments for the trials that were included in the analysis. Nine trials (including 3801 participants) were rated as low risk of bias the remainder (27 trials; 15,549 participants) were at high risk of bias—primarily due to lack of blinding.

### Mortality

We included 36 trials, with 19,350 patients and 5,269 deaths, in our network comparing tocilizumab and sarilumab, with or without corticosteroids, and corticosteroids in comparison with standard care or placebo (6, 26–28, 30, 31, 34, 35, 37–40, 42–46, 48). Nine trials could not be included in the analysis because they did not report outcome data, or, for trials that compared IL-6 receptor blockers with standard care or placebo, we could not retrieve subgroup data based on concomitant treatment with corticosteroids (29, 32, 33, 36, 41, 47). Figure 2 presents the network plot. Supplementary 1 contains data for the network meta-analysis.

**Figure 2:**
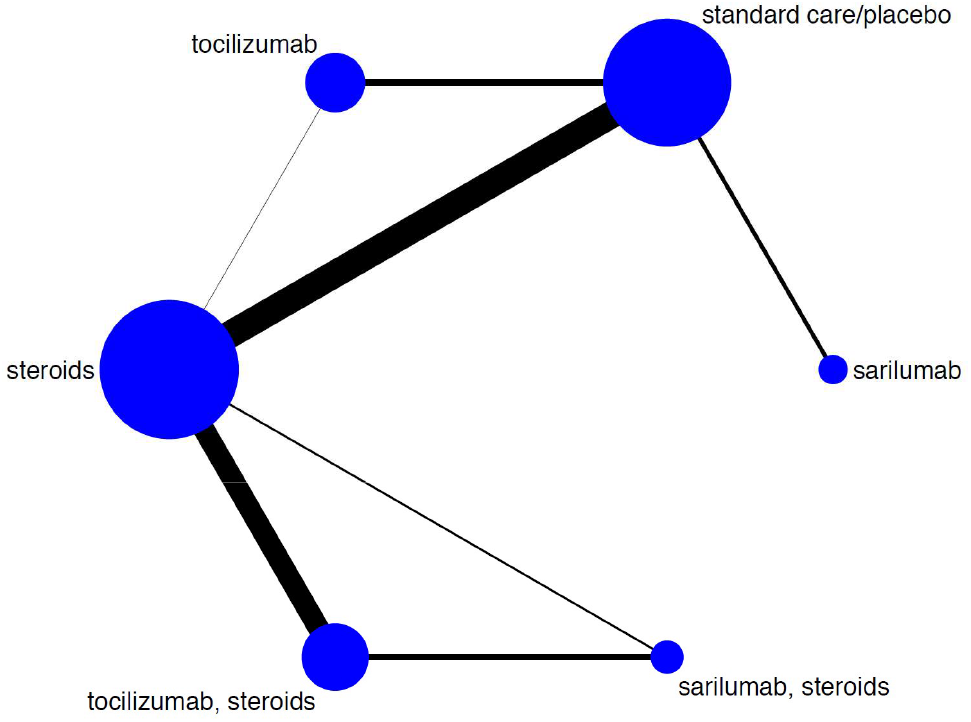
Network diagram. (nodes are weighted according to the number of studies evaluating each treatment and edges weighted according to the precision (inverse variance) of the direct estimate for each pairwise comparison).

**Figure 3:**
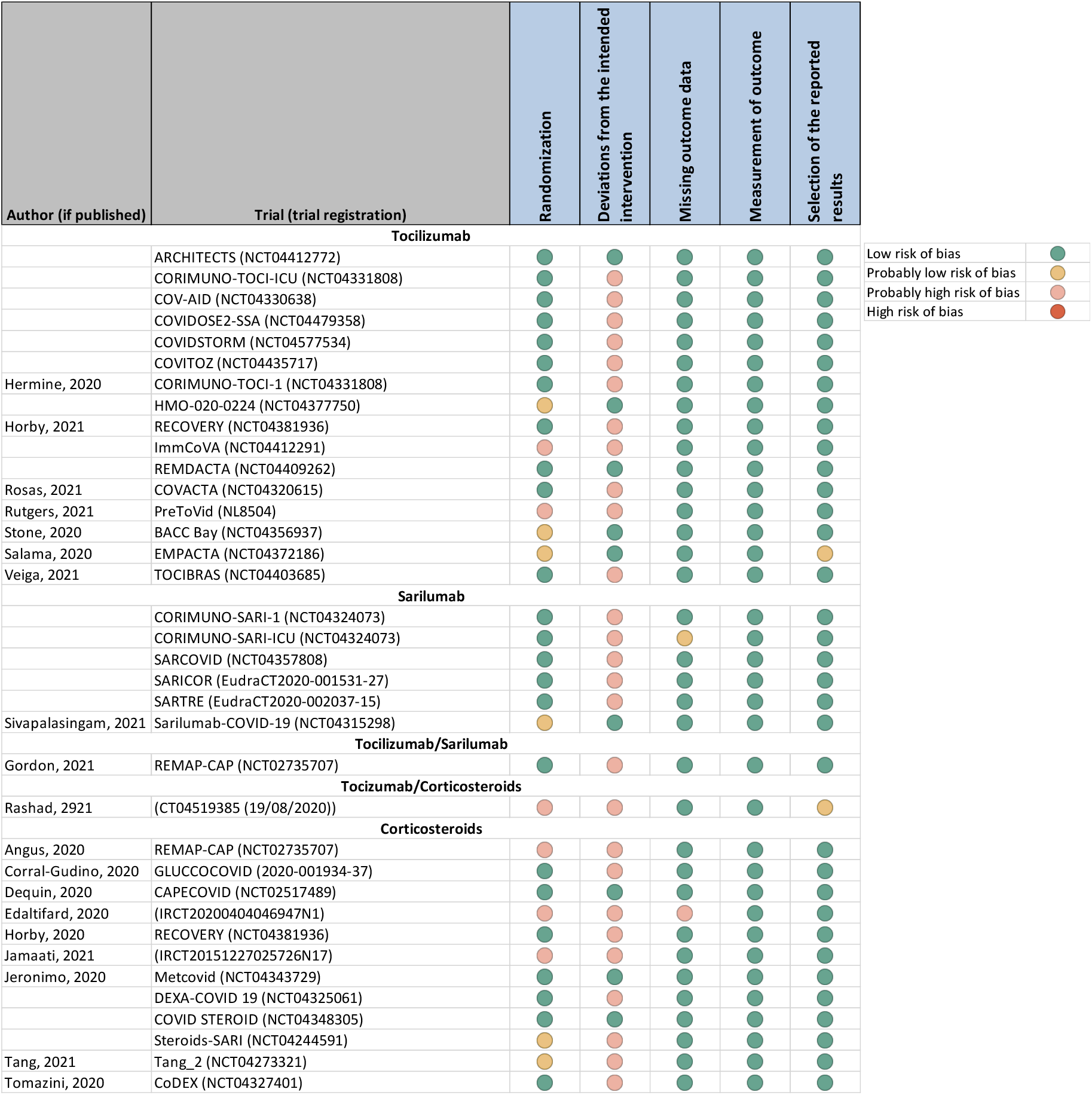
Risk of bias of trials included in the network meta-analysis.

Table 2 presents results from the network meta-analysis. Tocilizumab and sarilumab alone may not reduce mortality compared to standard care and corticosteroids probably reduce the risk of death. Compared to corticosteroids alone, tocilizumab in combination with corticosteroids probably reduces mortality and sarilumab in combination with corticosteroids may reduce mortality. In combination with corticosteroids, sarilumab may have similar effects to tocilizumab in reducing mortality. Supplementary 2 presents all direct and indirect comparisons and their certainty of evidence.

**Table 2:**
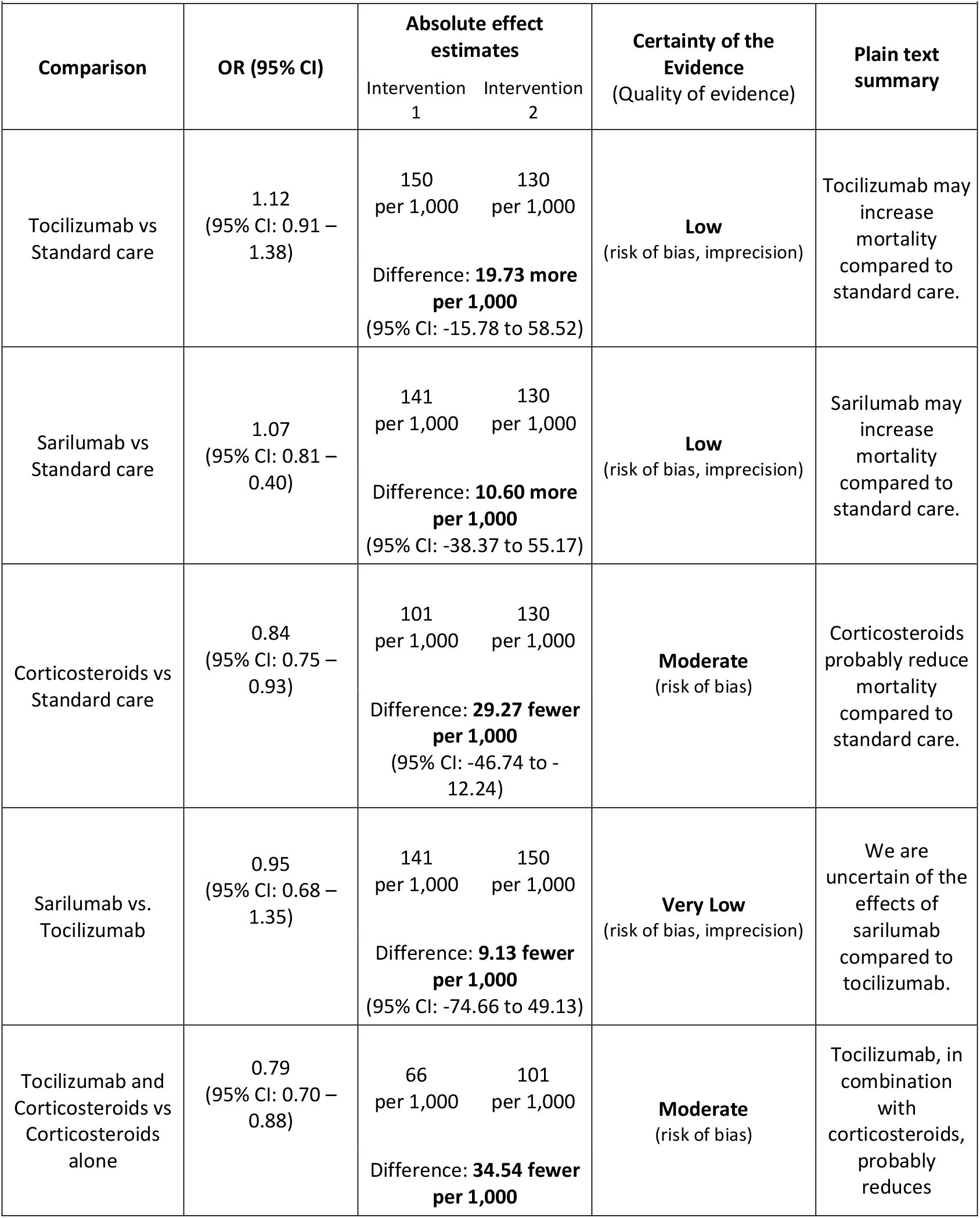
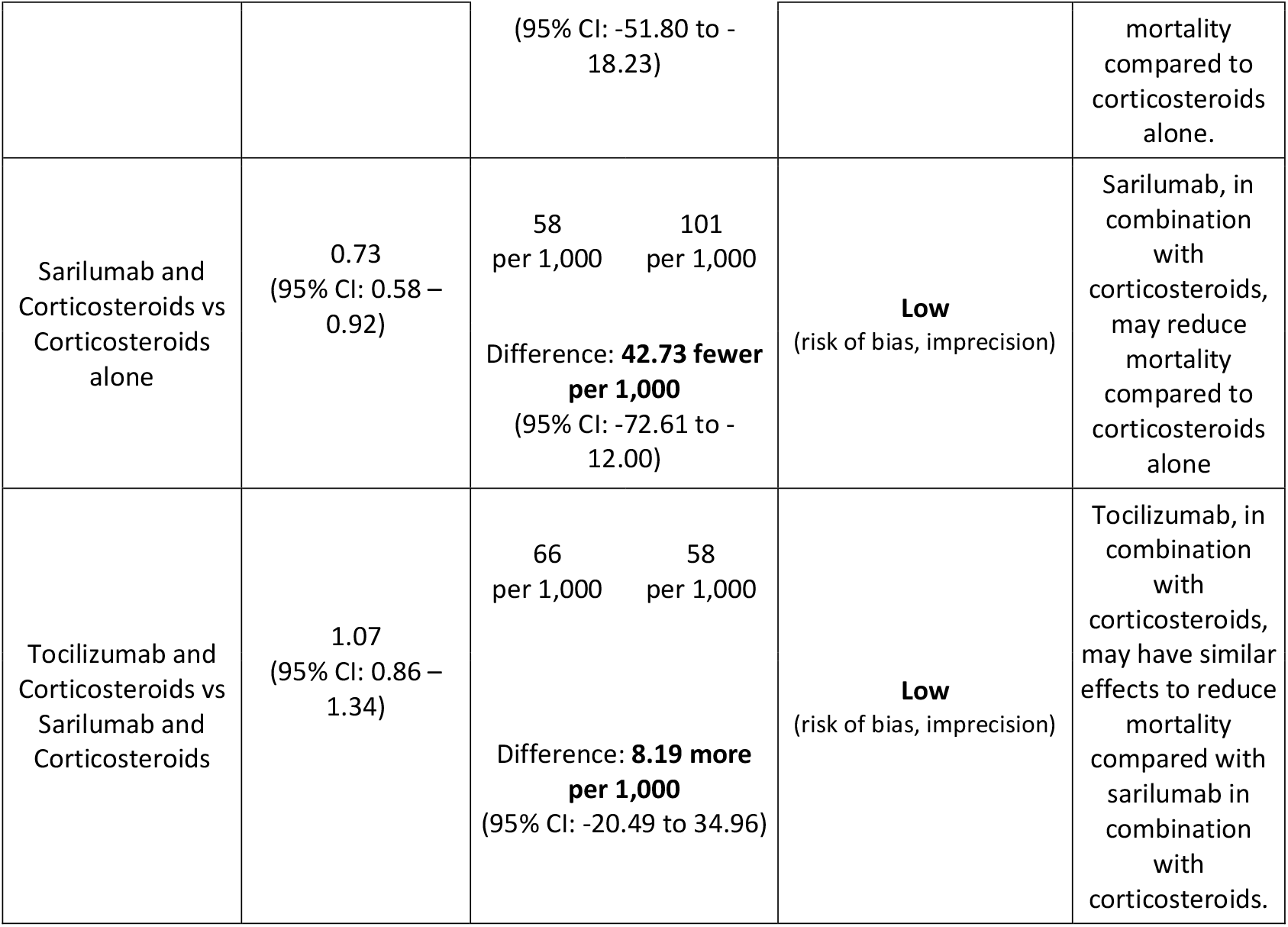
**Summary of findings for network meta-analysis comparing tocilizumab and sarilumab, alone or in combination with corticosteroids**

Supplementary 3 presents results from the random-effects model. Results from the random-effects model were largely consistent with results from the fixed-effects model—though the random-effects model produced effect estimates that were more imprecise due to incorporation of an additional heterogeneity parameter in the model.

## Discussion

### Main findings

This systematic review and network meta-analysis provides a comprehensive overview of the evidence for IL-6 receptor blockers, alone or in combination with corticosteroids. We show that IL-6 receptor blockers, when added to a standard care that includes corticosteroids, in patients with severe or critical COVID-19, probably reduce mortality. Whether or not IL-6 receptor blockers have any impact on mortality without concomitant use of corticosteroids remains uncertain. Sarilumab appears to have similar effectiveness as tocilizumab.

### Findings in context

Our findings are consistent with a prospective meta-analysis (11), and the largest trials on IL-6 receptor blockers, RECOVERY and REMAP-CAP (6, 7). Though several smaller trials did not find a benefit with tocilizumab, the totality of the evidence suggests that this is probably because smaller individual trials were underpowered to detect such a modest reduction in mortality.

Our study adds to the evidence base by showing that IL-6 receptor blockers may reduce mortality when added to a standard care regimen that includes corticosteroids and that sarilumab may have a similar effect on mortality as tocilizumab. This result is largely driven by the component of the REMAP-CAP trial that directly compared sarilumab to tocilizumab.

### Strengths and limitations

The strengths of this review include the comprehensive search and screening strategy. In addition to trials that we identified as part of our own search, we also included trials from two prospective meta-analyses that included an inception cohort of registered trials thereby minimizing the effects of publication bias (11, 15).

Our findings are limited by the risk of bias of the trials, the majority of which were at high risk of bias due to lack of blinding. Lack of blinding may introduce bias through differences in co-interventions between randomized groups. We took a conservative approach and rated down the certainty of evidence for risk of bias. Some, including the linked WHO guideline panel, did not consider lack of blinding to be a serious concern for mortality, because it is an objective outcome (13).

We only considered corticosteroid use at the time of randomization and some patients probably received corticosteroids after randomization, but were considered in this review not to have received concomitant corticosteroids. Whether or not IL-6 receptor blockers reduce mortality when not co-administered with corticosteroids remains uncertain.

Four trials that we included in our systematic review were only available as preprint publications. Including preprints in meta-analyses may increase the precision of estimates, allow timely dissemination, and may minimize the effects of publication bias. It may, however, reduce the credibility of evidence syntheses and risk serious errors if important differences appear in later published reports. As part of our living systematic review and network meta-analysis, we have been maintaining a comprehensive comparison of differences in key methods and results between preprints and publications. Differences between preprints and peer reviewed publications have mostly been limited to baseline patient characteristics and any changes we have observed so far would not have resulted in a meaningful change to the pooled effect estimates or certainty of evidence (2).

## Conclusion

Evidence from this systematic review and network meta-analysis shows that IL-6 receptor blockers when added to a standard care that includes corticosteroids, in patients with severe or critical COVID-19, probably reduce mortality. The available evidence suggests that tocilizumab and sarilumab may be similarly effective. Our findings support linked WHO guidelines on IL-6 receptor blockers, which recommends using either tocilizumab or sarilumab for patients with severe or critical COVID-19 (13).

## Data Availability

All data are included in the manuscript.

## References

1. COVID-19 Worldometer [Available from: https://www.worldometers.info/coronavirus/.

2. Siemieniuk RA, Bartoszko JJ, Ge L, et al. Drug treatments for covid-19: living systematic review and network meta-analysis. Bmj. 2020;370:m2980.

3. Pitre T, Jones A, Su J, et al. Inflammatory biomarkers as independent prognosticators of 28-day mortality for COVID-19 patients admitted to general medicine or ICU wards: a retrospective cohort study. Intern Emerg Med. 2021:1–10.

4. Chen N, Zhou M, Dong X, et al. Epidemiological and clinical characteristics of 99 cases of 2019 novel coronavirus pneumonia in Wuhan, China: a descriptive study. Lancet. 2020;395(10223):507–13.

5. Del Valle DM, Kim-Schulze S, Huang HH, et al. An inflammatory cytokine signature predicts COVID-19 severity and survival. Nat Med. 2020;26(10):1636–43.

6. Tocilizumab in patients admitted to hospital with COVID-19 (RECOVERY): a randomised, controlled, open-label, platform trial. Lancet. 2021;397(10285):1637–45.

7. Gordon AC, Mouncey PR, Al-Beidh F, et al. Interleukin-6 Receptor Antagonists in Critically Ill Patients with Covid-19. N Engl J Med. 2021;384(16):1491–502.

8. Stone JH, Frigault MJ, Serling-Boyd NJ, et al. Efficacy of Tocilizumab in Patients Hospitalized with Covid-19. N Engl J Med. 2020;383(24):2333–44.

9. Hermine O, Mariette X, Tharaux PL, et al. Effect of Tocilizumab vs Usual Care in Adults Hospitalized With COVID-19 and Moderate or Severe Pneumonia: A Randomized Clinical Trial. JAMA Intern Med. 2021;181(1):32–40.

10. Salvarani C, Dolci G, Massari M, et al. Effect of Tocilizumab vs Standard Care on Clinical Worsening in Patients Hospitalized With COVID-19 Pneumonia: A Randomized Clinical Trial. JAMA Intern Med. 2021;181(1):24–31.

11. Shankar-Hari M, Vale C, Godolphin P, et al. Association of administration of interleukin-6 antagonists with mortality and other outcomes among hospitalized patients with COVID-19: a prospective meta-analysis. JAMA. 2021.

12. Verma AA, Pai M, Saha S, et al. Managing drug shortages during a pandemic: tocilizumab and COVID-19. Cmaj. 2021;193(21):E771–e6.

13. Rochwerg B, Siemieniuk RA, Agoritsas T, et al. A living WHO guideline on drugs for covid-19. Bmj. 2020;370:m3379.

14. Marshall IJ, Noel-Storr A, Kuiper J, et al. Machine learning for identifying Randomized Controlled Trials: An evaluation and practitioner’s guide. Res Synth Methods. 2018;9(4):602–14.

15. Sterne JAC, Murthy S, Diaz JV, et al. Association Between Administration of Systemic Corticosteroids and Mortality Among Critically Ill Patients With COVID-19: A Meta-analysis. Jama. 2020;324(13):1330–41.

16. Sterne JAC, Savović J, Page MJ, et al. RoB 2: a revised tool for assessing risk of bias in randomised trials. Bmj. 2019;366:4898.

17. Turner RM, Jackson D, Wei Y, et al. Predictive distributions for between-study heterogeneity and simple methods for their application in Bayesian meta-analysis. Stat Med. 2015;34(6):984–98.

18. Chaimani A, Higgins JP, Mavridis D, et al. Graphical tools for network meta-analysis in STATA. PLoS One. 2013;8(10):e76654.

19. Centers for Disease Control and Prevention. COVIDView. A weekly surveillance summary of U.S COVID-19 activity 2020 [Available from: https://www.cdc.gov/coronavirus/2019-ncov/covid-data/covidview/index.html.

20. Centers for Disease Control and Prevention. Daily updates of totals by week and state: provisional death counts for coronavirus disease 2019 (COVID-19) 2020 [Available from: https://www.cdc.gov/nchs/nvss/vsrr/COVID19/index.htm.

21. ISARIC (International Severe Acute Respiratory and Emerging Infections Consortium). COVID-19 Report: 08 June 2020. medRxiv 2020.

22. Hultcrantz M, Rind D, Akl EA, et al. The GRADE Working Group clarifies the construct of certainty of evidence. J Clin Epidemiol. 2017;87:4–13.

23. Puhan MA, Schünemann HJ, Murad MH, et al. A GRADE Working Group approach for rating the quality of treatment effect estimates from network meta-analysis. Bmj. 2014;349:g5630.

24. Brignardello-Petersen R, Mustafa RA, Siemieniuk RAC, et al. GRADE approach to rate the certainty from a network meta-analysis: addressing incoherence. J Clin Epidemiol. 2019;108:77–85.

25. Brignardello-Petersen R, Bonner A, Alexander PE, et al. Advances in the GRADE approach to rate the certainty in estimates from a network meta-analysis. J Clin Epidemiol. 2018;93:36–44.

26. Hermine O, Mariette X, Tharaux P-L, et al. Effect of Tocilizumab vs Usual Care in Adults Hospitalized With COVID-19 and Moderate or Severe Pneumonia: A Randomized Clinical Trial. JAMA Internal Medicine. 2020.

27. Rosas IO, Bräu N, Waters M, et al. Tocilizumab in Hospitalized Patients with Severe Covid-19 Pneumonia. N Engl J Med. 2021.

28. Rutgers AaW, Peter E. and van der Holt, Bronno and Postma, Simone and van Vonderen, Marit G.A. and Piersma Djura P. and Postma, Douwe and van den Berge, Maarten and Jong, Eefje and de Vries, Marten and van der Burg, Leonie and Huugen, Dennis and van der Poel, Marjolein and Kampschreur Linda M. and Nijland, Marcel and Strijbos Jaap H. and Tamminga, Menno and Mutsaers Pim G. N. J. and Schol-Gelok, Suzanne and Dijkstra-Tiekstra, Margriet and Sidorenkov, Grigory and Vincenten, Julien and van Geffen, Wouter H. and Knoester, Marjolein and Kosterink, Jos and Gans, Reinold and Stegeman, Coen and Huls, Gerwin and van Meerten, Tom,. Timely Administration of Tocilizumab Improves Survival of Hospitalized COVID-19 Patients.. Available at SSRN: https://ssrncom/abstract=3834311 or http://dxdoiorg/102139/ssrn3834311. 2021.

29. Soin AS, Kumar K, Choudhary NS, et al. Tocilizumab plus standard care versus standard care in patients in India with moderate to severe COVID-19-associated cytokine release syndrome (COVINTOC): an open-label, multicentre, randomised, controlled, phase 3 trial. (2213–2619 (Electronic)).

30. Stone JH, Frigault MJ, Serling-Boyd NJ, et al. Efficacy of Tocilizumab in Patients Hospitalized with Covid-19. New England Journal of Medicine. 2020.

31. Salama C, Han J, Yau L, et al. Tocilizumab in Patients Hospitalized with Covid-19 Pneumonia. N Engl J Med. 2021;384(1):20–30.

32. Salvarani C, Dolci G, Massari M, et al. Effect of Tocilizumab vs Standard Care on Clinical Worsening in Patients Hospitalized With COVID-19 Pneumonia: A Randomized Clinical Trial. JAMA Internal Medicine. 2020.

33. Talaschian M, Akhtari M, Mahmoudi M, et al. Tocilizumab Failed to Reduce Mortality in Severe COVID-19 Patients: Results From a Randomized Controlled Clinical Trial. Respiratory Research. 2021.

34. Veiga VC, Prats J, Farias DLC, et al. Effect of tocilizumab on clinical outcomes at 15 days in patients with severe or critical coronavirus disease 2019: randomised controlled trial. BMJ. 2021;372:84.

35. Sivapalasingam S, Lederer DJ, Bhore R, et al. A Randomized Placebo-Controlled Trial of Sarilumab in Hospitalized Patients with Covid-19. medRxiv. 2021:2021.05.13.21256973.

36. Lescure FX, Honda H, Fowler RA, et al. Sarilumab in patients admitted to hospital with severe or critical COVID-19: a randomised, double-blind, placebo-controlled, phase 3 trial. The Lancet Respiratory medicine. 2021;9(5):522–32.

37. The Writing Committee for the REMAP-CAP Investigators. Effect of Hydrocortisone on Mortality and Organ Support in Patients With Severe COVID-19: The REMAP-CAP COVID-19 Corticosteroid Domain Randomized Clinical Trial. Jama. 2020;02:02.

38. Corral-Gudino L, Bahamonde A, Arnaiz-Revillas F, et al. Methylprednisolone in adults hospitalized with COVID-19 pneumonia : An open-label randomized trial (GLUCOCOVID). Wiener klinische Wochenschrift. 2021:1–9.

39. Dequin PF, Heming N, Meziani F, et al. Effect of Hydrocortisone on 21-Day Mortality or Respiratory Support Among Critically Ill Patients With COVID-19: A Randomized Clinical Trial. Jama. 2020;02:02.

40. Edalatifard M, Akhtari M, Salehi M, et al. Intravenous methylprednisolone pulse as a treatment for hospitalised severe COVID-19 patients: results from a randomised controlled clinical trial. The European respiratory journal. 2020.

41. Farahani RH MR, Nezami-Asl A et al. Evaluation of the Efficacy of Methylprednisolone Pulse Therapy in Treatment of Covid-19 Adult Patients with Severe Respiratory Failure: Randomized, Clinical Trial,. 09 September 2020, PREPRINT (Version 1) available at Research Square [+https://doiorg/1021203/rs3rs-66909/v1+]. 2020.

42. The RECOVERY Collaborative Group. Dexamethasone in Hospitalized Patients with Covid-19. N Engl J Med. 2021;384(8):693–704.

43. Jamaati H, Hashemian SM, Farzanegan B, et al. No clinical benefit of high dose corticosteroid administration in patients with COVID-19: A preliminary report of a randomized clinical trial. Eur J Pharmacol. 2021;897:173947.

44. Jeronimo CMP, Farias MEL, Val FFA, et al. Methylprednisolone as Adjunctive Therapy for Patients Hospitalized With COVID-19 (Metcovid): A Randomised, Double-Blind, Phase IIb, Placebo-Controlled Trial. Clinical infectious diseases : an official publication of the Infectious Diseases Society of America. 2020.

45. Tang X, Feng YM, Ni JX, et al. Early Use of Corticosteroid May Prolong SARS-CoV-2 Shedding in Non-Intensive Care Unit Patients with COVID-19 Pneumonia: A Multicenter, Single-Blind, Randomized Control Trial. Respiration; international review of thoracic diseases. 2021:1–11.

46. Tomazini BM, Maia IS, Cavalcanti AB, et al. Effect of Dexamethasone on Days Alive and Ventilator-Free in Patients With Moderate or Severe Acute Respiratory Distress Syndrome and COVID-19: The CoDEX Randomized Clinical Trial. Jama. 2020;02:02.

47. Vaira LA, Hopkins C, Petrocelli M, et al. Efficacy of corticosteroid therapy in the treatment of long-lasting olfactory disorders in COVID-19 patients. Rhinology. 2021;59(1):21–5.

48. Rashad A, Mousa S, Nafady-Hego H, et al. Short term survival of critically ill COVID-19 Egyptian patients on assisted ventilation treated by either Dexamethasone or Tocilizumab. Sci Rep. 2021;11(1):8816.

49. The WHO Rapid Evidence Appraisal for COVID-19 Therapies (REACT) Working Group. Association Between Administration of Systemic Corticosteroids and Mortality Among Critically Ill Patients With COVID-19: A Meta-analysis. Jama. 2020;02:02.

50. Gordon AC, Mouncey PR, Al-Beidh F, et al. Interleukin-6 Receptor Antagonists in Critically Ill Patients with Covid-19. N Engl J Med. 2021.

