## Supplementary 1 for "Tocilizumab and sarilumab alone or in combination with corticosteroids for COVID-19: A systematic review and network meta-analysis"

| | | | | Dichotomous outcomes | | | | |
|---|---|---|---|---|---|---|---|---|
| Ref ID | 1st Author | Intervention name | Subgroup | Outcome<br>1=Mortality [closest to 90 days]<br>2=Mechanical ventilation (includes both invasive and non-invasive) [closest to 90 days]<br>3=Admission to hospital [within 28 days]<br>4=Adverse effects leading to discontinuation [within 28 days]<br>5=Viral clearance [closest to 7 days +/- 3 days]<br>6=Venous thromboembolism [only for anticoagulants]<br>7=Clinically important bleeding [only for | Follow-up time (days) | N analyzed | Number of events | Comments |
| 5743, 2967: | Corral-Gudino | thylprednisol | NA | 1 | 28 | 35 | 7 | NA |
| 5743, 2967: | Corral-Gudino | standard care | NA | 1 | 28 | 29 | 5 | NA |
| 9, 7002, 33 | Horby_1 | examethason | NA | 1 | 28 | 2104 | 482 | NA |
| 9, 7002, 33 | Horby_1 | standard care | NA | 1 | 28 | 4321 | 1110 | NA |
| 12062 | Dequin | ıydrocortison | NA | 1 | 21 | 75 | 11 | NA |
| 12062 | Dequin | placebo | NA | 1 | 21 | 73 | 20 | NA |
| 12141 | Tomazini | examethason | NA | 1 | 28 | 151 | 85 | NA |
| 12141 | Tomazini | standard care | NA | 1 | 28 | 148 | 91 | NA |
| 12146 | Angus | ortisone (fixe | NA | 1 | NR | 137 | 41 | NA |
| 12146 | Angus | sone (shock-c | NA | 1 | NR | 141 | 37 | NA |
| 12146 | Angus | standard care | NA | 1 | NR | 101 | 33 | NA |
| r Patients \ | Sterne | examethason | NA | 1 | 28 | 7 | 2 | NA |
| r Patients \ | Sterne | standard care | NA | 1 | 28 | 12 | 2 | NA |
| D-19 and S | Sterne | hydrocortis one | NA | 1 | 28 | 15 | 6 | NA |
| D-19 and S | Sterne | placebo | NA | 1 | 28 | 14 | 2 | NA |
| Ill Patients | Sterne | thylprednisol | NA | 1 | 30 | 24 | 13 | NA |
| Ill Patients | Sterne | standard care | NA | 1 | 30 | 23 | 13 | NA |
| 13208 | Edalatifard | thylprednisol | NA | 1 | NR | 34 | 2 | NA |
| 13208 | Edalatifard | standard care | NA | 1 | NR | 28 | 12 | NA |
| 2 | Farahani | dnisolone, pr | NA | 1 | NR | 14 | 0 | NA |
| 2 | Farahani | standard care | NA | 1 | NR | 15 | NR | NA |
| 27640 | Tang_2 | thylprednisol | NA | 1 | NR | 43 | 0 | NA |
| 27640 | Tang_2 | standard care | NA | 1 | NR | 43 | 1 | NA |
| 31583 | Jamaati | examethason | NA | 1 | 28 | 25 | 16 | NA |
| 31583 | Jamaati | standard care | NA | 1 | 28 | 25 | 15 | NA |
| 10699 | Jeronimo | thylprednisol | NA | 1 | 28 | 209 | 79 | NA |
| 10699 | Jeronimo | placebo | NA | 1 | 28 | 207 | 80 | NA |
| :osas, 3330 | COVACTA_NCT04320615 | tocilizumab | Steroids | 1 | 90 | 56 | 16 | NA |
| :osas, 3330 | COVACTA_NCT04320615 | placebo | Steroids | 1 | 90 | 41 | 14 | NA |
| :osas, 3330 | COVACTA_NCT04320615 | tocilizumab | No steroids | 1 | 90 | 238 | 56 | NA |
| :osas, 3330 | COVACTA_NCT04320615 | placebo | No steroids | 1 | 90 | 103 | 22 | NA |
| 30368 | RECOVERY_NCT04381936 | tocilizumab | Steroids | 1 | 28 | 1664 | 482 | NA |
| 30368 | RECOVERY_NCT04381936 | standard care | Steroids | 1 | 28 | 1721 | 600 | NA |
| 30368 | RECOVERY_NCT04381936 | tocilizumab | No steroids | 1 | 28 | 357 | 139 | NA |
| 30368 | RECOVERY_NCT04381936 | standard care | No steroids | 1 | 28 | 367 | 127 | NA |
| :CTS_NCT0 | ARCHITECTS_NCT04412772 | tocilizumab | Steroids | 1 | 90 | 9 | 0 | NA |
| :CTS_NCT0 | ARCHITECTS_NCT04412772 | placebo | Steroids | 1 | 90 | 11 | 2 | NA |
| :CTS_NCT0 | ARCHITECTS_NCT04412772 | tocilizumab | No steroids | 1 | 90 | 1 | 0 | NA |
| :CTS_NCT0 | ARCHITECTS_NCT04412772 | placebo | No steroids | 1 | 90 | 0 | 0 | NA |
| 16283 | BACC Bay_NCT04356937 | tocilizumab | Steroids | 1 | 28 | 3 | 0 | NA |
| 16283 | BACC Bay_NCT04356937 | placebo | Steroids | 1 | 28 | 1 | 0 | NA |
| 16283 | BACC Bay_NCT04356937 | tocilizumab | No steroids | 1 | 28 | 158 | 9 | NA |
| 16283 | BACC Bay_NCT04356937 | placebo | No steroids | 1 | 28 | 81 | 4 | NA |
| 16128 | ORIMUNO-TOCI-1_NCT0433180 | tocilizumab | Steroids | 1 | 90 | 10 | 1 | NA |
| 16128 | ORIMUNO-TOCI-1_NCT0433180 | standard care | Steroids | 1 | 90 | 12 | 4 | NA |
| 16128 | ORIMUNO-TOCI-1_NCT0433180 | tocilizumab | No steroids | 1 | 90 | 53 | 6 | NA |
| 16128 | ORIMUNO-TOCI-1_NCT0433180 | standard care | No steroids | 1 | 90 | 55 | 7 | NA |
| 16127 | )RIMUNO-TOCI-ICU_NCT043318 | tocilizumab | Steroids | 1 | 90 | 8 | 4 | NA |
| 16127 | )RIMUNO-TOCI-ICU_NCT043318 | standard care | Steroids | 1 | 90 | 4 | 2 | NA |
| 16127 | )RIMUNO-TOCI-ICU_NCT043318 | tocilizumab | No steroids | 1 | 90 | 41 | 8 | NA |
| 16127 | )RIMUNO-TOCI-ICU_NCT043318 | standard care | No steroids | 1 | 90 | 39 | 11 | NA |
| ID_NCT043 | COV-AID_NCT04330638 | tocilizumab | Steroids | 1 | 90 | 48 | 7 | NA |
| ID_NCT043 | COV-AID_NCT04330638 | standard care | Steroids | 1 | 90 | 42 | 3 | NA |
| ID_NCT043 | COV-AID_NCT04330638 | tocilizumab | No steroids | 1 | 90 | 33 | 3 | NA |
| ID_NCT043 | COV-AID_NCT04330638 | standard care | No steroids | 1 | 90 | 30 | 6 | NA |
| :2-SSA_NCT | COVIDOSE2-SSA_NCT04479358 | tocilizumab | Steroids | 1 | 28 | 6 | 0 | NA |
| :2-SSA_NCT | COVIDOSE2-SSA_NCT04479358 | standard care | Steroids | 1 | 28 | 2 | 1 | NA |
| :2-SSA_NCT | COVIDOSE2-SSA_NCT04479358 | tocilizumab | No steroids | 1 | 28 | 13 | 0 | NA |

| | | | | | | | | |
|---|---|---|---|---|---|---|---|---|
| 2-SSA_NCT | COVIDOSE2-SSA_NCT04479358 | standard care | No steroids | 1 | 28 | 6 | 1 | NA |
| 6340, 2364 | EMPACTA_NCT04372186 | tocilizumab | Steroids | 1 | 90 | 200 | 27 | NA |
| 6340, 2364 | EMPACTA_NCT04372186 | placebo | Steroids | 1 | 90 | 112 | 15 | NA |
| 6340, 2364 | EMPACTA_NCT04372186 | tocilizumab | No steroids | 1 | 90 | 49 | 2 | NA |
| 6340, 2364 | EMPACTA_NCT04372186 | placebo | No steroids | 1 | 90 | 16 | 0 | NA |
| -0224_NCT | HMO-020-0224_NCT04377750 | tocilizumab | Steroids | 1 | 90 | 31 | 16 | NA |
| -0224_NCT | HMO-020-0224_NCT04377750 | placebo | Steroids | 1 | 90 | 15 | 9 | NA |
| -0224_NCT | HMO-020-0224_NCT04377750 | tocilizumab | No steroids | 1 | 90 | 6 | 2 | NA |
| -0224_NCT | HMO-020-0224_NCT04377750 | placebo | No steroids | 1 | 90 | 2 | 1 | NA |
| 1, Gordon_ | AP-CAP_1_NCT02735707_tociliz | tocilizumab | Steroids | 1 | 90 | 214 | 63 | NA |
| 1, Gordon_ | AP-CAP_1_NCT02735707_tociliz | standard care | Steroids | 1 | 90 | 217 | 80 | NA |
| 1, Gordon_ | AP-CAP_1_NCT02735707_sarilu | sarilumab | Steroids | 1 | 90 | 44 | 8 | NA |
| 1, Gordon_ | AP-CAP_1_NCT02735707_sarilu | standard care | Steroids | 1 | 90 | 52 | 13 | NA |
| 1, Gordon_ | AP-CAP_1_NCT02735707_tociliz | tocilizumab | No steroids | 1 | 90 | 127 | 33 | NA |
| 1, Gordon_ | AP-CAP_1_NCT02735707_tociliz | standard care | No steroids | 1 | 90 | 129 | 44 | NA |
| 1, Gordon_ | AP-CAP_1_NCT02735707_sarilu | sarilumab | No steroids | 1 | 90 | 4 | 2 | NA |
| 1, Gordon_ | AP-CAP_1_NCT02735707_sarilu | standard care | No steroids | 1 | 90 | 11 | 5 | NA |
| Gordon_2 | REMAP-CAP_2_NCT02735707 | tocilizumab | Steroids | 1 | NR | 589 | 215 | NA |
| Gordon_2 | REMAP-CAP_2_NCT02735707 | sarilumab | Steroids | 1 | NR | 431 | 146 | NA |
| CTA_NCT0 | REMDACTA_NCT04409262 | tocilizumab | Steroids | 1 | 28 | 358 | 69 | NA |
| CTA_NCT0 | REMDACTA_NCT04409262 | placebo | Steroids | 1 | 28 | 181 | 39 | NA |
| CTA_NCT0 | REMDACTA_NCT04409262 | tocilizumab | No steroids | 1 | 28 | 72 | 9 | NA |
| CTA_NCT0 | REMDACTA_NCT04409262 | placebo | No steroids | 1 | 28 | 29 | 2 | NA |
| 27470 | TOCIBRAS_NCT04403685 | tocilizumab | Steroids | 1 | 90 | 31 | 9 | NA |
| 27470 | TOCIBRAS_NCT04403685 | standard care | Steroids | 1 | 90 | 34 | 5 | NA |
| 27470 | TOCIBRAS_NCT04403685 | tocilizumab | No steroids | 1 | 90 | 34 | 8 | NA |
| 27470 | TOCIBRAS_NCT04403685 | standard care | No steroids | 1 | 90 | 30 | 2 | NA |
| -SARI-1_N | ORIMUNO-SARI-1_NCT0432407 | sarilumab | Steroids | 1 | 90 | 4 | 2 | NA |
| -SARI-1_N | ORIMUNO-SARI-1_NCT0432407 | standard care | Steroids | 1 | 90 | 3 | 0 | NA |
| -SARI-1_N | ORIMUNO-SARI-1_NCT0432407 | sarilumab | No steroids | 1 | 90 | 72 | 14 | NA |
| -SARI-1_N | ORIMUNO-SARI-1_NCT0432407 | standard care | No steroids | 1 | 90 | 65 | 10 | NA |
| SARI-ICU_N | RIMUNO-SARI-ICU_NCT043240 | sarilumab | Steroids | 1 | 90 | 0 | 0 | NA |
| SARI-ICU_N | RIMUNO-SARI-ICU_NCT043240 | standard care | Steroids | 1 | 90 | 2 | 0 | NA |
| SARI-ICU_N | RIMUNO-SARI-ICU_NCT043240 | sarilumab | No steroids | 1 | 90 | 48 | 14 | NA |
| SARI-ICU_N | RIMUNO-SARI-ICU_NCT043240 | standard care | No steroids | 1 | 90 | 31 | 13 | NA |
| VID_NCT04 | SARCOVID_NCT04357808 | sarilumab | Steroids | 1 | 90 | 17 | 1 | NA |
| VID_NCT04 | SARCOVID_NCT04357808 | standard care | Steroids | 1 | 90 | 8 | 0 | NA |
| VID_NCT04 | SARCOVID_NCT04357808 | sarilumab | No steroids | 1 | 90 | 3 | 1 | NA |
| VID_NCT04 | SARCOVID_NCT04357808 | standard care | No steroids | 1 | 90 | 2 | 0 | NA |
| udraCT202 | ARICOR_EudraCT2020-001531-2 | sarilumab | Steroids | 1 | 28 | 66 | 2 | NA |
| udraCT202 | ARICOR_EudraCT2020-001531-2 | standard care | Steroids | 1 | 28 | 34 | 3 | NA |
| udraCT202 | ARICOR_EudraCT2020-001531-2 | sarilumab | No steroids | 1 | 28 | 10 | 1 | NA |
| udraCT202 | ARICOR_EudraCT2020-001531-2 | standard care | No steroids | 1 | 28 | 5 | 1 | NA |
| draCT2020 | ARTRE_EudraCT2020-002037-1 | sarilumab | Steroids | 1 | 28 | 70 | 2 | NA |
| draCT2020 | ARTRE_EudraCT2020-002037-1 | standard care | Steroids | 1 | 28 | 70 | 1 | NA |
| draCT2020 | ARTRE_EudraCT2020-002037-1 | sarilumab | No steroids | 1 | 28 | 0 | 0 | NA |
| draCT2020 | ARTRE_EudraCT2020-002037-1 | standard care | No steroids | 1 | 28 | 0 | 0 | NA |
| OVIDSTORI | COVIDSTORM | tocilizumab | Steroids | 1 | 90 | 13 | 1 | NA |
| OVIDSTORI | COVIDSTORM | standard care | Steroids | 1 | 90 | 17 | 1 | NA |
| OVIDSTORI | COVIDSTORM | tocilizumab | No steroids | 1 | 90 | 0 | 0 | NA |
| OVIDSTORI | COVIDSTORM | standard care | No steroids | 1 | 90 | 9 | 0 | NA |
| ImmCoVA | ImmCoVA | tocilizumab | Steroids | 1 | 28 | 21 | 2 | NA |
| ImmCoVA | ImmCoVA | standard care | Steroids | 1 | 28 | 26 | 2 | NA |
| ImmCoVA | ImmCoVA | tocilizumab | No steroids | 1 | 28 | 1 | 0 | NA |
| ImmCoVA | ImmCoVA | standard care | No steroids | 1 | 28 | 1 | 0 | NA |
| Rutgers | PreToVid_Trial NL8504 | tocilizumab | Steroids | 1 | 28 | 159 | 20 | NA |
| Rutgers | PreToVid_Trial NL8504 | standard care | Steroids | 1 | 28 | 167 | 32 | NA |
| Rutgers | PreToVid_Trial NL8504 | tocilizumab | No steroids | 1 | 28 | 11 | 1 | NA |
| Rutgers | PreToVid_Trial NL8504 | standard care | No steroids | 1 | 28 | 11 | 2 | NA |
| 41237 | Rashad | tocilizumab | NA | 1 | 14 | 74 | 60 | NA |
| 41237 | Rashad | examethason | NA | 1 | 14 | 75 | 45 | NA |
| vapalasinga | REGENERON-P2_NCT04315298 | sarilumab | Steroids | 1 | 90 | 62 | 28 | NA |
| vapalasinga | REGENERON-P2_NCT04315298 | standard care | Steroids | 1 | 90 | 14 | 6 | NA |
| vapalasinga | REGENERON-P2_NCT04315298 | sarilumab | No steroids | 1 | 90 | 305 | 81 | NA |
| vapalasinga | REGENERON-P2_NCT04315298 | standard care | No steroids | 1 | 90 | 76 | 18 | NA |
| vapalasinga | REGENERON-P3_NCT04315298 | sarilumab | Steroids | 1 | 90 | 345 | 99 | NA |
| vapalasinga | REGENERON-P3_NCT04315299 | standard care | Steroids | 1 | 90 | 99 | 34 | NA |
| vapalasinga | REGENERON-P3_NCT04315298 | sarilumab | No steroids | 1 | 90 | 699 | 198 | NA |

| | | | | | | | | |
|---|---|---|---|---|---|---|---|---|
| vapalasinga | REGENERON-P3_NCT04315299 | standard care | No steroids | 1 | 90 | 187 | 50 | NA |
| COVITOZ | COVITOZ | tocilizumab | Steroids | 1 | 90 | 6 | 0 | NA |
| COVITOZ | COVITOZ | standard care | Steroids | 1 | 90 | 6 | 0 | NA |
| COVITOZ | COVITOZ | tocilizumab | No steroids | 1 | 90 | 4 | 0 | NA |
| COVITOZ | COVITOZ | standard care | No steroids | 1 | 90 | 3 | 0 | NA |
