## Supplementary 2 for "Tocilizumab and sarilumab alone or in combination with corticosteroids for COVID-19: A systematic review and network meta-analysis"

| Comparison | | Direct estimate | | | Indirect estimate | | | Network estimate | | | | | | | | |
|---|---|---|---|---|---|---|---|---|---|---|---|---|---|---|---|---|
| | | Relative estimate | | | Relative estimate | | | Relative estimate | | | Absolute estimate (per 1,000) | | | | | Reasons |
| Treatment 1 | Treatment 2 | oint estimat | CI lower limi | CI upper limi | oint estimat | CI lower limi | CI upper limi | Point estim | CI lower lim | CI upper lin | Point estim | CI lower lim | CI upper lin | Final rating | Higher certainty estimate | |
| sarilumab | sarilumab, steroids | NA | NA | NA | 1.75 | 1.21 | 2.56 | 1.75 | 1.21 | 2.56 | 82.6 | 31.26 | 125 | LOW | NMA | RoB, Imprecision |
| sarilumab | standard care/placebo | 1.05 | 0.68 | 1.59 | NA | NA | NA | 1.07 | 0.81 | 1.4 | 10.6 | -38.37 | 55.17 | LOW | NMA | RoB, Imprecision |
| sarilumab | steroids | NA | NA | NA | 1.28 | 0.95 | 1.72 | 1.28 | 0.95 | 1.72 | 39.87 | -7.76 | 82.29 | LOW | NMA | RoB, Imprecision |
| sarilumab | tocilizumab | NA | NA | NA | 0.95 | 0.68 | 1.35 | 0.95 | 0.68 | 1.35 | -9.13 | -74.66 | 49.13 | VERY LOW | NMA | RoB, Imprecisionx2 |
| sarilumab | tocilizumab, steroids | NA | NA | NA | 1.63 | 1.2 | 2.25 | 1.63 | 1.2 | 2.25 | 74.41 | 29.82 | 112.66 | LOW | NMA | RoB, Imprecision |
| sarilumab, steroids | standard care/placebo | NA | NA | NA | 0.61 | 0.47 | 0.79 | 0.61 | 0.47 | 0.79 | -72 | -106.11 | -37.34 | MODERATE | NMA | RoB |
| sarilumab, steroids | steroids | 0.82 | 0.49 | 1.41 | 0.69 | 0.52 | 0.92 | 0.73 | 0.58 | 0.92 | -42.73 | -72.61 | -12 | LOW | NMA | RoB, Imprecision |
| sarilumab, steroids | tocilizumab | NA | NA | NA | 0.54 | 0.39 | 0.76 | 0.54 | 0.39 | 0.76 | -91.73 | -144.09 | -42.48 | MODERATE | NMA | RoB |
| tocilizumab, steroids | sarilumab, steroids | 1.12 | 0.34 | 3.7 | 0.95 | 0.63 | 1.42 | 1.07 | 0.86 | 1.34 | 8.19 | -20.49 | 34.96 | LOW | NMA | RoB, Imprecision |
| steroids | standard care/placebo | 0.84 | 0.64 | 1.07 | 0.36 | 0.16 | 0.77 | 0.84 | 0.75 | 0.93 | -29.27 | -46.74 | -12.24 | MODERATE | NMA | RoB |
| tocilizumab | standard care/placebo | 0.98 | 0.7 | 1.36 | 2.47 | 1.17 | 5.4 | 1.12 | 0.91 | 1.38 | 19.73 | -15.78 | 58.52 | LOW | NMA | RoB, Imprecision |
| standard care/placebo | tocilizumab, steroids | NA | NA | NA | 1.54 | 1.31 | 1.79 | 1.54 | 1.31 | 1.79 | 63.81 | 40.66 | 88.06 | MODERATE | NMA | RoB |
| tocilizumab | steroids | 2.86 | 0.75 | 10.95 | 1.23 | 0.97 | 1.57 | 1.34 | 1.07 | 1.68 | 49 | 10.17 | 91.51 | MODERATE | NMA | RoB |
| tocilizumab, steroids | steroids | 0.8 | 0.64 | 1.01 | 0.91 | 0.57 | 1.46 | 0.79 | 0.7 | 0.88 | -34.54 | -51.8 | -18.23 | MODERATE | NMA | RoB |
| tocilizumab | tocilizumab, steroids | NA | NA | NA | 1.72 | 1.32 | 2.23 | 1.72 | 1.32 | 2.23 | 83.54 | 41.39 | 129.99 | MODERATE | NMA | RoB |
