## Supplementary 3 for "Tocilizumab and sarilumab alone or in combination with corticosteroids for COVID-19: A systematic review and network meta-analysis"

| | | NMA estimate, odds ratio | | | NMA estimate, risk difference | | | Indirect estimate, odds ratio | | | Direct estimate, odds ratio | | |
|---|---|---|---|---|---|---|---|---|---|---|---|---|---|
| **Treatment 1** | **Treatment 2** | **OR** | **LCL** | **UCL** | **RD** | **LCL** | **UCL** | **OR** | **LCL** | **UCL** | **OR** | **LCL** | **UCL** |
| sarilumab | sarilumab, steroids | 1.75 | 1.07 | 2.92 | 82.4 | 12.1 | 136.9 | | | | | | |
| sarilumab | standard care/placebo | 1.06 | 0.76 | 1.49 | 9.1 | -52.1 | 62.6 | | | | 1.05 | 0.68 | 1.59 |
| sarilumab | steroids | 1.34 | 0.92 | 2.01 | 46.7 | -14.6 | 100.4 | | | | | | |
| **sarilumab** | **tocilizumab** | **0.97** | **0.64** | **1.48** | -8.2 | -88.4 | 61.3 | | | | | | |
| sarilumab | tocilizumab, steroids | 1.67 | 1.09 | 2.59 | 76.2 | 15.0 | 126.7 | | | | | | |
| sarilumab, steroids | standard care/placebo | 0.61 | 0.41 | 0.87 | -73.3 | -121.2 | -22.5 | | | | | | |
| **sarilumab, steroids** | **steroids** | **0.76** | **0.56** | **1.06** | -35.7 | -74.2 | 8.1 | 0.71 | 0.44 | 1.15 | 0.82 | 0.49 | 1.41 |
| sarilumab, steroids | tocilizumab | 0.55 | 0.36 | 0.85 | -90.6 | -158.0 | -26.4 | | | | | | |
| **tocilizumab, steroids** | **sarilumab, steroids** | **1.05** | **0.77** | **1.43** | 6.2 | -36.0 | 43.4 | 0.96 | 0.59 | 1.57 | 1.12 | 0.34 | 3.70 |
| steroids | standard care/placebo | 0.79 | 0.64 | 0.95 | -37.5 | -70.3 | -8.2 | 0.35 | 0.15 | 0.80 | 0.84 | 0.64 | 1.07 |
| tocilizumab | standard care/placebo | 1.09 | 0.85 | 1.40 | 17.3 | -25.7 | 65.5 | 2.42 | 1.07 | 5.59 | 0.98 | 0.70 | 1.36 |
| standard care/placebo | tocilizumab, steroids | 1.57 | 1.20 | 2.08 | 67.1 | 29.0 | 106.9 | | | | | | |
| tocilizumab | steroids | 1.39 | 1.03 | 1.90 | 54.9 | 5.9 | 111.4 | 1.23 | 0.89 | 1.70 | 2.86 | 0.75 | 10.95 |
| **tocilizumab, steroids** | **steroids** | **0.80** | **0.67** | **0.99** | -29.6 | -55.2 | -2.2 | 0.92 | 0.50 | 1.74 | 0.80 | 0.64 | 1.01 |
| tocilizumab | tocilizumab, steroids | 1.72 | 1.21 | 2.46 | 84.4 | 29.5 | 146.1 | | | | | | |

OR, odds ratio; LCL, 95% lower credible limit; UCL, 95% upper credible limit
